# Attributable is Preventable: Corrected and revised estimates of population attributable fraction of TB related to undernutrition in 30 high TB burden countries

**DOI:** 10.1101/2021.12.09.21267540

**Authors:** Anurag Bhargava, Madhavi Bhargava, Andrea Beneditti, Anura Kurpad

## Abstract

**Introduction:** The Global TB Report 2020 estimated the population attributable fractions (PAF) for the major risk factors of TB. Undernourishment emerged as the leading risk factor accounting for 19% of the cases. The WHO however used the terms undernourishment and undernutrition interchangeably in its computation of PAF. Undernourishment is an indirect model derived estimate of decreased per capita energy availability, while undernutrition is defined by direct anthropometric measurements of nutritional status.

**Methods:** We re-estimated the PAF of undernutrition (instead of undernourishment) in 30 high TB burden countries, using the prevalence of undernutrition (age standardized estimate of BMI < 18.5 kg/m^2^ in adults for both sexes), and the relative risk (RR) of 3.2. Further, we revised PAF estimates of undernutrition with an RR of 4.49, in light of recent evidence.

**Findings:** Twenty four percent of TB in high burden countries is attributable to undernutrition. The PAF of undernutrition was highest in Asian countries, unlike the PAF of undernourishment that was highest in Africa. The corrected estimate led up to 65% increase in number of cases attributable to undernutrition in Asian countries. More than one-third to nearly half of TB cases in India could be attributable to undernutrition.

**Interpretation:** Estimation of the PAF of TB related to undernutrition is methodologically valid and operationally relevant, rather than PAF related to undernourishment. Addressing undernutrition, the leading driver of TB in high TB burden countries (especially Asia) could enable achievement of END TB milestones of TB incidence for 2025.

## Background

Tuberculosis (TB) remains one of the major public health problems that cause morbidity and premature mortality in the low and middle income countries (LMIC). In the year 2019, an estimated 10 million people fell ill with TB, 1.2 million HIV negative patients with TB died, and 0.21 million patient with HIV-TB co-infection died [1]. Ninety-five percent of the new cases as well as deaths occurred in the LMICs [1]. The new END TB strategy has ambitious goals for reduction of TB incidence and mortality and end the epidemic by 2035. The strategy has milestones for 2020 and 2025 that require TB incidence to decline by 20% and 50%, and TB mortality to decline by 35% and 75% by 2020 and 2025 respectively compared to the baseline 2015 [2]. The current rate of decline of 9% of new cases and 14% of mortality since 2015 is not adequate to reach these milestones [1]. The END TB strategy envisages public health interventions to address the social determinants and risk factors of TB to complement the biomedical interventions, and achieve the 10% annual decline in TB incidence required to reach the 2025 milestone [2].

Infection with mycobacterium tuberculosis is necessary but not sufficient to result in TB disease. Latent infection of TB (LTBI) is prevalent in nearly one-quarter of the world population but only 10% of this progress to active TB in their life time; innate and adaptive immunity prevents the progression in the rest [3,4]. Conditions like HIV infection, undernutrition (defined for example by a low body mass index, BMI), diabetes, alcohol use disorders and tobacco use, impair immune responses, increase the risk of active TB and act as drivers of the TB epidemic at the population level [5,6]. The strong, consistent and inverse association of BMI and TB incidence has been noted in large historical cohorts [7], as well as recent cohort studies which have adjusted for confounders [8-10]. A systematic review of cohort studies concluded that this association of BMI and TB was causal [7].

The population attributable fraction (PAF) is an epidemiologic measure useful in the estimation of the public health impact of a particular risk factor for an outcome. It is the proportion by which the incidence an outcome in the entire population would be reduced if the risk factor was eliminated [11]. The key parameters that determine the PAF are the prevalence of a risk factor and the relative risk (RR) of disease associated with it [11].

An estimate of the PAFs of the aforementioned risk factors in 22 high TB burden countries was first reported more than a decade ago [12]. These were 11.0% for HIV, 26.9% for undernutrition, 7.5% for diabetes, 9.8% for alcohol use disorders, and 15.8% for smoking [12]. While undernutrition accounted for the largest proportion of cases, the major risk factors driving the epidemic varied in individual countries. In South Africa, the PAF for HIV was 69.1% due to high HIV prevalence (18.1%), where as it was 35% for alcohol in Russia. The leading risk factor was undernutrition accounting for 31.6% of TB cases in India [12]. For estimation of the PAF of undernutrition, the prevalence of *undernourishment* (PoU) in the total population, according to Food and Agriculture Organization (FAO) estimates of 2008 were used, instead of prevalence of *undernutrition* [12]. The RRs of the risk factors in the above estimation were 26.7 for HIV, 3.2 for undernutrition, 3.1 for diabetes, 2.9 for alcohol use disorders, and 2.0 for smoking [12]. The RR of 3.2 (95% CI 3.1–3.3) for undernutrition was based on a meta-analysis that compared the risk of TB for body-mass index (BMI) 16 kg/m^2^ versus 25 kg/m^2^, based on an average reduction in TB incidence of 13.8% (95% CI 13.4–14.2) per unit increase in BMI [7]. This systematic review included six cohort studies, the most recent of which had not published final results at that time [7].

Recently the Global TB Report of 2020 updated the current PAFs for the five risk factors based on the global data; 19% for undernourishment, 7.7% for HIV, 3.1% for diabetes, 8.1% for alcohol use disorders, and 7.1% for smoking. The number of attributable cases related to these risk factors were estimated as 2.2 million for undernourishment, 0.76 million for HIV, 0.35 million for diabetes, 0.72 million for alcohol use disorders, and 0.70 million for smoking.

*Undernourishment* emerged as the leading risk factor, accounting for nearly three times the cases attributable to HIV, or that attributable to the combined diabetes, alcohol and tobacco [1].

### Rationale for correction of population attributable fraction of “undernourishment” as reported in the Global TB report 2020

The PAF in 2010 was reported for the risk factor of *undernutrition*, while in 2020 it was reported for the risk factor of *undernourishment* [1, 12]. These two terms have been used interchangeably in both the reports. However, undernourishment as defined by FAO is different from undernutrition for public health and clinical purposes [13].

The FAO measure of undernourishment is derived by comparing the usual food consumption expressed in terms of dietary energy (kcal) with energy requirement norms. The part of the population with food consumption below this norm is considered undernourished and PoU estimates the percentage of population that have “habitual insufficient food consumption to provide, on an average, the amount of dietary energy required for maintaining a normal, active and healthy life”[14]. It is therefore a measure of chronic food insecurity, not based on actual energy intakes but per capita energy availability [13]. These estimates use three sources of data; the food balance sheets for the country, inequality in energy intakes, and the country energy requirements by sex and age group [14]. These are used to derive the four parameters in the analytic formula of PoU (Box-1) [14]. Thus PoU is not based on any direct individual measurements but is an indirect national-level model-based indicator that measures the level of chronic hunger in a population. PoU estimates have a serious limitation as they do not allow disaggregated analysis to identify specific vulnerable populations to be identified in a country. It is also not possible to compute margins of error for the PoU.

#### Box-1

**Estimation of Prevalence of Undernourishment (PoU) by Food and Agriculture Organization (FAO)**

PoU estimates are obtained using the following analytic formula [14]:

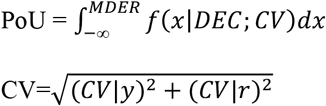

Dietary Energy Consumption (DEC): It is an estimate of the per capita level of the average habitual, daily dietary energy consumption in the population.

MDER (Minimum Dietary Energy Requirement): It is the amount of energy needed for light activity and a minimum acceptable weight for attained height, and depending upon the gender and age structure of the population, it varies by country and from year to year.

Coefficient of Variation (CV): It is an estimate of the CV of the distribution of per capita levels of habitual dietary energy consumption and is the combination of two components. The *CV*|*y* is the component that is associated with differences in energy requirements in the population of a country. *CV*|*r* is the part of the variation that can be associated with differences in the socio-economic characteristics of the households

Undernutrition, on the other hand, is a physiological condition based on actual measurement of individual nutritional status [13]. It “describes the status of persons whose heights and weights lie below the lower limits of the ranges established for healthy people”[15]. In adults, undernutrition is defined based on a low BMI (<18.5kg/m^2^) that reflects low body energy stores or chronic energy deficiency [16]. It has also been accepted as a criterion for clinical diagnosis of malnutrition/undernutrition in a consensus statement [5].

Thus undernourishment and undernutrition are two temporally distinct terms derived from different analytical approaches; undernourishment precedes the potential development of undernutrition, and these are not interchangeable. The FAO considered them complementary measures (Figure 1) [13]. Previous studies have highlighted that there may be differences in estimates of undernourishment and undernutrition determined by anthropometry as the former refers to poor availability of food while the latter reflects the outcome of undernourishment, and also affected by repeated infections, poor care and neglect [17].

**Figure 1:**
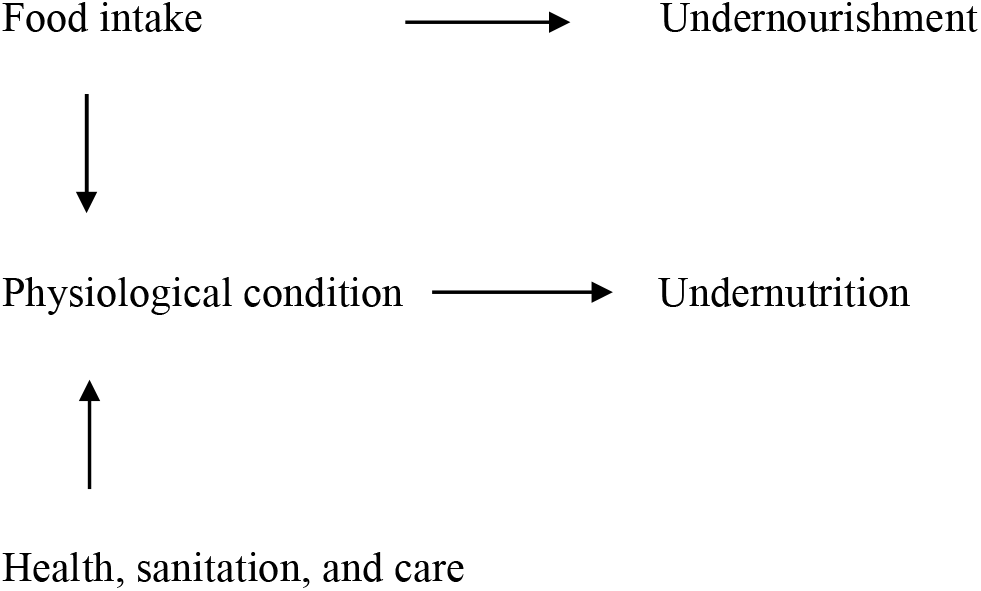
Differentiating undernourishment and undernutrition.

Further, an estimate of PAF for any risk factor relies on the prevalence as well as the RR of the same exposure or risk factor, and this is not the case in the present methodology. In the case of the estimation of PAF of TB due to undernourishment the prevalence used is based on a measurement of chronic food insecurity while the RR is based on an anthropometric measurement. The 2010 estimates of PAF for undernutrition err in using the PoU for its computation. On the other hand, the 2020 estimate of PAF for undernourishment errs in using the RR of undernutrition. It may be pointed out that RR for undernourishment is not available.

### Recent evidence in favor of revising RR of TB related to undernutrition

Previous reviews of tuberculosis epidemiology have suggested a point estimate of the relative risk (RR) of undernutrition as 4·0 with a range of 2.0 to 6.0 [18]. The RR of 3.2 was derived from a particular systematic review of cohort studies in 2009 [7]. The results of studies in the past decade suggest a need for an upward revision of the relative risk of undernutrition for TB. The final results of a US-based cohort study included in the systematic review were published in 2012 [10]. This cohort study involved a nationally representative sample of participants, while others in the meta-analysis were often restricted to males, or special populations like the elderly [7]. It also controlled for multiple confounders to arrive at the effects of nutritional status on TB incidence, and one of the few which had risk estimates for those with BMI <18.5 kg/m^2^ [10]. The hazard ratio for the development of TB in those with BMI <18.5 kg/m^2^ was 12.43 (95% CI: 5.75, 26.95) while the RR for development of TB was 4.49 (95% CI: 2.28 - 8.86, p<0.0005) compared to those with normal BMI (18.5-25 kg/m^2^) [10]. A modeling study estimating the impact of reducing undernutrition on TB incidence in some high TB burden states in India [19], incorporated the final published results of this study into those of the previous meta-analysis [7]. Using a penalized spline model to estimate the RR for different quartiles of BMI, it estimated RRs of 4.95 and 3.00 for those with BMI quartiles of ≤17.8 kg/m^2^, 17.80-≤19.64kg/m^2^ compared to the reference of 22.25 kg/m^2^ thus providing a continuum of risk [19]. Another limitation of the cohort studies in the systematic review was that they were all conducted in high income countries [7]. Recently in a cohort of household contacts of patients with active pulmonary tuberculosis in India, HIV and undernutrition were the only 2 independent risk factors associated with the risk of incident TB [20]. The adjusted incidence rate ratio (aIRR) of undernutrition was 6.16 (95% CI: 1.89, 20.03) measured as a composite index of low BMI (< 18.5 kg/m^2^) in adults, and less than two z-scores of weight or BMI for age in children [20]. Taken together these results suggest that the RR of 3.2 based on a comparison of BMI of 16 kg/m^2^ vs. BMI of 25 kg/m^2^ may be a conservative estimate, since the risk and rate ratio of those with BMI less than 18.5 were exceeding 4.0 in recent studies [10,19,20].

## Objectives

The objective of the present study was to correct the estimates of PAF of undernutrition for TB using the prevalence and RR of undernutrition in 30 high burden countries. We further revised the estimate of PAF of undernutrition for TB using a revised RR based on recent evidence.

## Methods

The current estimated PAFs and PoU were taken from the Global TB Report 2020 [1]. Undernutrition was defined as a low BMI (<18.5 kg/m^2^) in adults [16]. For the prevalence of undernutrition, we used the age-standardized estimate of prevalence of underweight (BMI<18.5 kg/m^2^) in both sexes in 30 high TB burden countries from the BMI database of the WHO Global Health Observatory [21]. PAFs were derived using the Levin formula [22]:

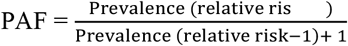

First, we derived a corrected estimate of PAF of undernutrition for TB by substituting the PoU with prevalence of undernutrition. As a next step, we further revised the PAF using RR for undernutrition as 4.49. Confidence intervals (CI) for the PAFs were estimated using the method suggested by Natarajan et al [23]. We further derived the number of cases attributable to undernutrition by multiplying the annual number of incident TB cases from the Global TB Report 2020 by the estimated PAFs (Figure 2).

**Figure 2:**
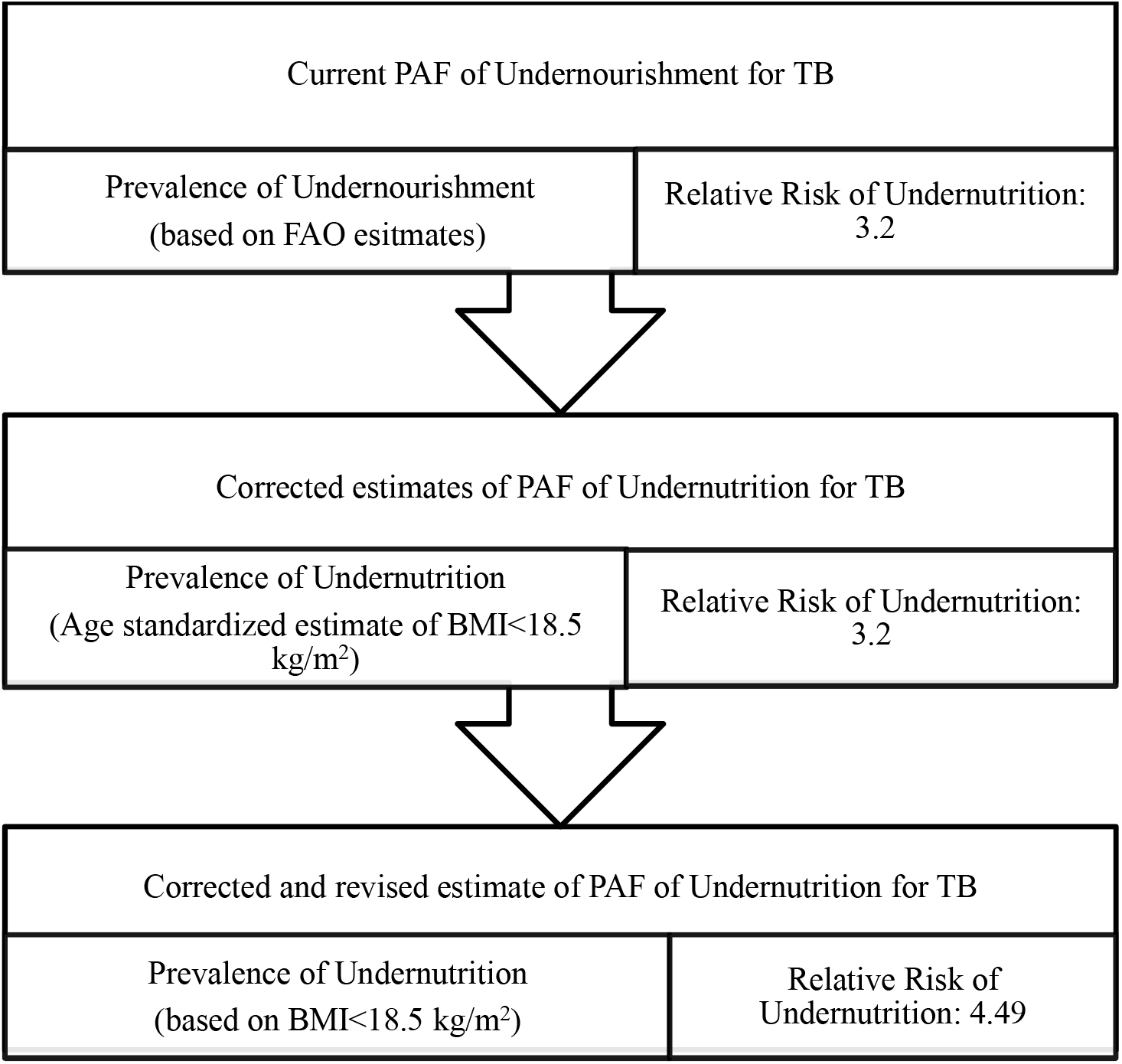
Steps in arriving corrected and revised estimates of Population Attributable Fraction of undernutrition for TB.

## Results

Table 1 shows the TB incidence and PoU in 30 high TB burden countries, the estimated PAF using a RR of 3.2 and the number of cases attributable to undernourishment. There were 8.62 million incident cases in these 30 countries. The median (IQR) of the PoU was 18% (10.6, 29.5), the median (IQR) of the PAF related to undernourishment was 28.3% (18.9, 39.3) and an estimated 23.6% (1.96 million) of incident cases in 28 high burden countries were attributable to undernourishment (excluding Democratic Republic of Congo and Papua New Guinea as they do not have data on PoU). In Asian countries, the PoU ranged from 7.8% in Thailand to 20% in Pakistan, while in African countries it ranged from 6·2% in South Africa to 60% in Central African Republic. The PAFs of undernourishment in African countries ranged from 12% in South Africa to 56% in Central African Republic, and in Asian countries from 14% in Thailand to 30% in Pakistan. The three countries with the highest PAFs of undernourishment globally were Central African Republic, Zimbabwe, and Democratic Republic of Korea; and overall the PAFs were higher in the African countries than the Asian countries.

**Table 1:**
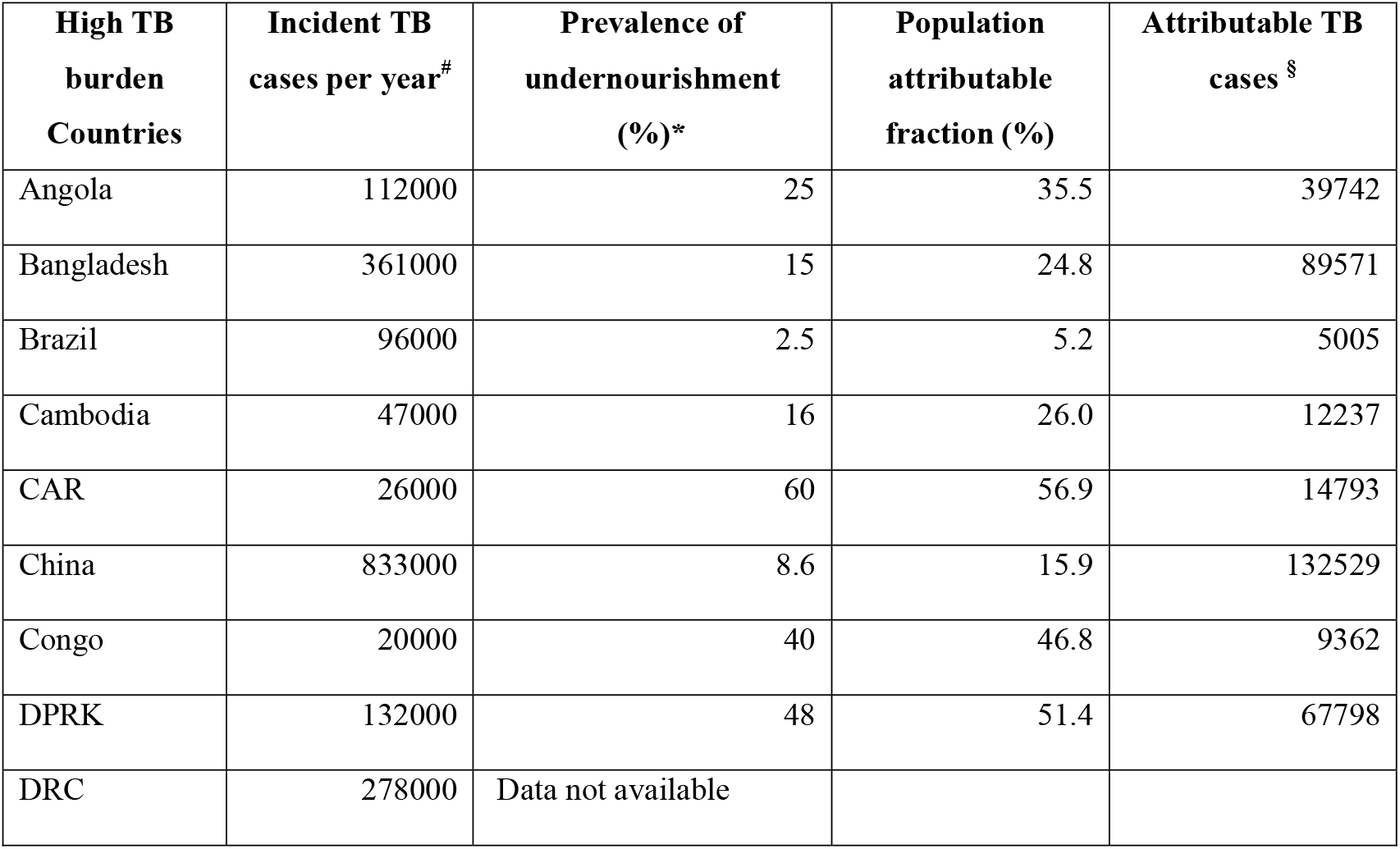
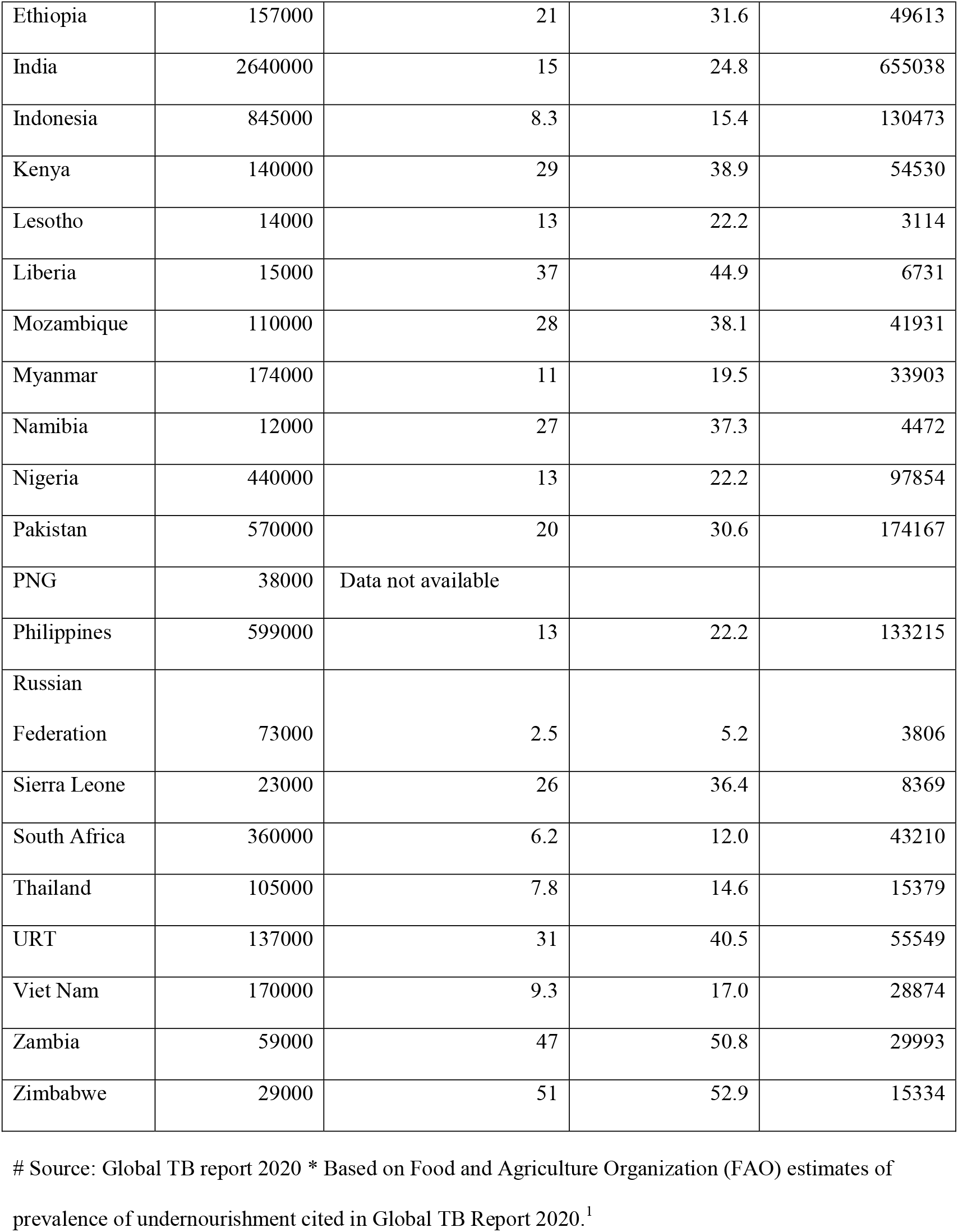
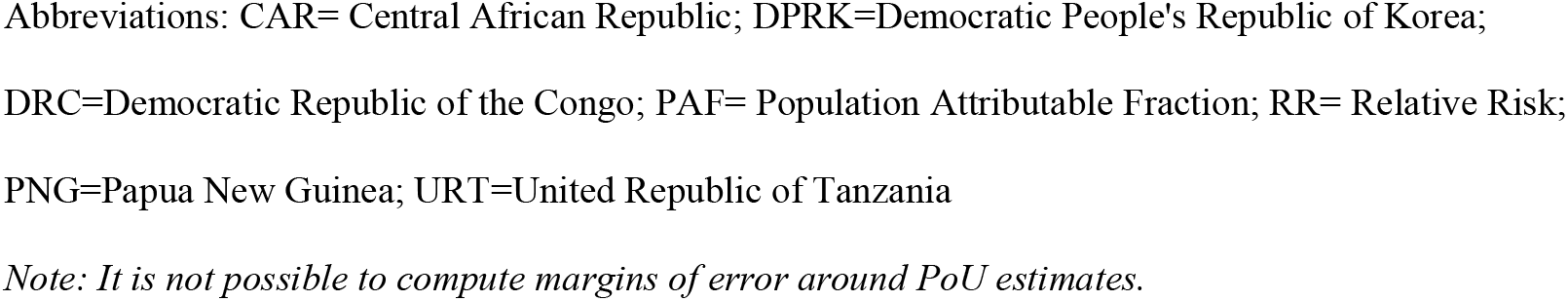
Population Attributable Fraction for Tuberculosis and attributable TB cases in high burden countries using prevalence of undernourishment (FAO estimates) and RR=3.2.

Table 2 shows the prevalence of undernutrition and the corrected estimates of PAF and attributable cases. The median (IQR) of prevalence of undernutrition was 11.2% (8.0, 14.4), the median (IQR) of the PAF related to undernutrition was 19.7% (15.0, 24.0), and 24% (2.07 million) of incident cases in all the 30 high burden countries were attributable to undernutrition. The prevalence of undernutrition was higher in the Asian countries than the PoU with a few exceptions and ranged from 12.3% in Philippines to 23.6% in India (the highest globally). The prevalence of undernutrition was higher than PoU by 10%, 57 %, and 90% in Thailand, India and Vietnam respectively. In the African countries, the prevalence of undernutrition was lower than PoU without exception and ranged from 4.7% in South Africa to the highest of 15.9% in Democratic Republic of Congo. In South Africa and Zimbabwe it was lower by 24% and 84% than the PoU estimates respectively. When the PAFs were recomputed replacing undernourishment with undernutrition, the median (IQR) PAF was 19.7 %(15.0, 24.0) with a range of 3% in Russia to 34.2% in India. The PAFs of undernutrition in African countries ranged from 9% in South Africa to 26% in Democratic Republic of Congo and in Asian countries from 10% in China to 34% in India. The three countries with the highest PAF of undernutrition globally were India, Bangladesh and Vietnam. Among the African countries, the number of attributable cases declined by a median (IQR) of 50% (35, 57). In the Asian countries the overall change was a median (IQR) increase in attributable cases by 16.7% (−10, 35.7) [data not in table]. The overall increase in attributable cases in Myanmar, Bangladesh, India, Indonesia and Vietnam was 25%, 29%, 38%, 43%, and 65% respectively. The number of cases attributable to undernutrition in Asian countries was 1.7 millions and in African countries about 0.37 millions.

**Table 2:** Population Attributable Fraction for Tuberculosis related to Undernutrition and attributable TB cases in high burden countries using prevalence of undernutrition based on anthropometry* and Relative Risk of 3.2 (95% CI: 3.1, 3.3)

| High TB burden Countries | Incident TB cases | Prevalence of undernutrition based on anthropometry [%,(95% CIs)] * | PAF for undernutrition<sup>#</sup> [%,(95% CIs)] | Attributable TB cases [(95% CIs)]<sup>§</sup> |
| --- | --- | --- | --- | --- |
| Angola | 112000 | 13.5 (7.2, 21.10) | 22.9 (10.4, 33.2) | 25,647 (11,643, 37,169) |
| Bangladesh | 361000 | 21.5 (16.80, 26.70) | 32.1 (24.9, 38.6) | 115,922 (89,725, 139,401) |
| Brazil | 96000 | 27.0 (18.0, 39.0) | 5.6 (3, 8.3) | 5,383 (2,915, 7,954) |
| Cambodia | 47000 | 13.6 (8.90, 19.10) | 23 (14, 31) | 10,824 (6,561, 14,587) |
| CAR | 26000 | 14.8 (8.9, 21.6) | 24.6 (13.6, 33.8) | 6,386 (3,537, 8,792) |
| China | 833000 | 5.3 (3.9, 6.9) | 10.4 (7, 14) | 86,985 (58,014, 116,427) |
| Congo | 20000 | 13.1 (8.9, 19.1) | 22.4 (13.2, 30.5) | 4,474 (2,636, 6,096) |
| DPRK | 132000 | 6.6 (3.1, 12.2) | 12.7 (2.8, 21.5) | 16,736 (3,749, 28,335) |
| DRC | 278000 | 15.9 (10.0, 22.5) | 25.9 (15.5, 34.8) | 72,044 (42,969, 96,746) |
| Ethiopia | 157000 | 15.8 (10.5, 21.9) | 25.8 (16.2, 34.1) | 40,497 (25,494, 53,498) |
| India | 2640000 | 23.6 (19.9, 27.6) | 34.2 (28.6, 39.3) | 384,898 (755,413, 1,038,631) |
| Indonesia | 845000 | 12.9 (9.3, 16.9) | 22.1 (15.2, 28.5) | 186,798 (128,076, 241,175) |
| Kenya | 140000 | 11.8 (7.6, 16.9) | 20.6 (11.9, 28.4) | 28,854 (16,696, 39,745) |
| Lesotho | 14000 | 7.9 (4.9, 11.6) | 14.8 (7.8, 21.4) | 2,073 (1,097, 2,991) |
| Liberia | 15000 | 8.3 (4.9, 12.50) | 15.4 (7.6, 22.7) | 2,316 (1,145, 3,398) |
| Mozambique | 110000 | 11.1 (7.1, 15.9) | 19.6 (11.2, 27.2) | 21,590 (12,374, 29,918) |
| Myanmar | 174000 | 14.6 (10.4, 19.5) | 24.3 (16.4, 31.4) | 42,302 (28,545, 54,711) |
| Namibia | 12000 | 9.7 (6.1, 14.1) | 17.6 (9.7, 24.8) | 2,110 (1,161, 2,981) |
| Nigeria | 440000 | 10.3 (7.3, 13.9) | 18.5 (12, 24.6) | 81,285 (52,777, 108,162) |
| Pakistan | 570000 | 15.0 (10.8, 19.9) | 24.8 (17, 31.9) | 141,429 (96,812, 181,693) |
| PNG | 38000 | 2.3 (1.0, 4.4) | 4.8 (0.7, 8.9) | 1,830 (281, 3,400) |
| Philippines | 599000 | 12.3 (8.4, 16.7) | 21.3 (13.6, 28.3) | 127,569 (81,626, 169,541) |
| Russian Federation | 73000 | 1.8 (1.1, 2.6) | 3.8 (1.9, 5.8) | 2,781 (1,409, 4,233) |
| Sierra Leone | 23000 | 10.3 (6.4, 14.9) | 18.5 (10.2, 26) | 4,249 (2,347, 5,977) |
| South Africa | 360000 | 4.7 (3.2, 6.4) | 9.4 (5.7, 13.1) | 33,736 (20,360, 47,290) |
| Thailand | 105000 | 8.6 (5.6, 12.3) | 15.9 (9.1, 22.4) | 16,705 (9,512, 23,472) |
| URT | 137000 | 11.0 (7.5, 15.1) | 19.5 (12.2, 26.2) | 26,694 (16,717, 35,923) |
| Viet Nam | 170000 | 17.7 (13.6, 22.1) | 28 (21.1, 34.3) | 47,645 (35,943, 58,342) |
| Zambia | 59000 | 11.2 (7.0, 16.0) | 19.8 (11.2, 27.5) | 11,664 (6,623, 16,202) |
| Zimbabwe | 29000 | 7.9 (5.0, 11.4) | 14.8 (8.1, 21.1) | 4,294 (2,360, 6,124) |

\*Source for age standardised BMI<18.5 kg/m<sup>2</sup> in both sexes reported in Global Health Observatory database on BMI (accessible at [https://www.who.int/data/gho/data/themes/topics/topic-](https://www.who.int/data/gho/data/themes/topics/topic-details/GHO/body-mass-index) [details/GHO/body-mass-index](https://www.who.int/data/gho/data/themes/topics/topic-details/GHO/body-mass-index)); #PAF calculated using a RR of 3.2(95% CI: 3.1-3.3); §Attributable cases using prevalence of undernutrition and RR of 3.2.; Abbreviations: CAR= Central African Republic; DPRK=Democratic People's Republic of Korea; DRC=Democratic Republic of the Congo; PAF= Population Attributable Fraction; RR= Relative Risk; PNG=Papua New Guinea; URT=United Republic of Tanzania

Table 3 shows the PAFs corrected using prevalence of undernutrition and revised RR of 4.49 (95% CI: 2.28, 8.86). The median (IQR) PAF attributable to undernutrition in 30 high TB burden countries was 28% (21.8, 33.4), and 32.9% (2.84 million) of incident cases were attributable to undernutrition. The revised PAF of undernutrition ranged from 5.9% in Russia to a maximum of 44.5% in India. The estimated number of cases attributable to undernutrition computed with a higher RR of undernutrition was 2.3 millions in Asia and 0.45 millions in Africa.

**Table 3:** Population Attributable Fraction for Tuberculosis related to undernutrition and attributable cases in high burden countries using prevalence of undernutrition based on anthropometry and Relative Risk of 4.49 (2.28, 8.86)

| High TB burden Countries | Incident TB cases | Prevalence of undernutrition based on anthropometry *[% , (95% CIs)] | PAF with revised RR<sup>s</sup> [% , (95% CIs)] | Attributable TB cases due to undernutrition estimated with revised RR <sup>#</sup>[(95% CIs)] |
| --- | --- | --- | --- | --- |
| Angola | 112000 | 13.5 (7.2, 21.10) | 32.0 (5.6,65.2) | 35869 (6271,73067) |
| Bangladesh | 361000 | 21.5 (16.80,26.70) | 42.9 (14.5,70.4) | 154756 (52210,254096) |
| Brazil | 96000 | 2.7 (1.80,3.90) | 8.6 (1.6,2.5) | 8267 (1513,24430) |
| Cambodia | 47000 | 13.6 (8.90,19.10) | 32.2 (7.7,63.0) | 15128 (3600,29595) |
| CAR | 26000 | 14.8 (8.9,21.6) | 34.1 (7.5,65.9) | 8855 (1937,17128) |
| China | 833000 | 5.3 (3.9,6.9) | 15.6 (3.7,38.0) | 130029 (30702,316860) |
| Congo | 20000 | 13.1 (8.9,19.1) | 31.4 (7.2,62.4) | 6275 (1440,12472) |
| DPRK | 132000 | 6.6 (3.1,12.2) | 18.7 (1.5,50.8) | 24713 (1943,67064) |
| DRC | 278000 | 15.9 (10.0,22.5) | 35.7 (8.6,66.6) | 99212 (23761,185848) |
| Ethiopia | 157000 | 15.8 (10.5,21.9) | 35.5 (9.0,66.1) | 55802 (14156,103834) |
| India | 2640000 | 23.6 (19.9,27.6) | 45.2 (17.0,71.0) | 1192346 (448963,1874920) |
| Indonesia | 845000 | 12.9 (9.3,16.9) | 31.0 (8.4,60.2) | 262326 (70712,508230) |
| Kenya | 140000 | 11.8 (7.6,16.9) | 29.2 (6.5,60.0) | 40837 (9063,83952) |
| Lesotho | 14000 | 7.9 (4.9,11.6) | 21.6 (4.2,50.7) | 3026 (583,7092) |
| Liberia | 15000 | 8.3 (4.9,12.50) | 22.5 (4.1,52.5) | 3369 (608,7879) |
| Mozambique | 110000 | 11.1 (7.1,15.9) | 27.9 (6.1,58.5) | 30714 (6694,64387) |
| Myanmar | 174000 | 14.6 (10.4,19.5) | 33.8 (9.1,63.4) | 58733 (15864,110332) |
| Namibia | 12000 | 9.7 (6.1,14.1) | 25.3 (5.2,55.5) | 3035 (623,6664) |
| Nigeria | 440000 | 10.3 (7.3,13.9) | 26.4 (6.5,55.2) | 116344 (28660,242829) |
| Pakistan | 570000 | 15.0 (10.8,19.9) | 34.4 (9.5,63.9) | 195862 (53971,364071) |
| PNG | 38000 | 20.3 (1.0,4.4) | 7.4 (0.04,27.1) | 2824 (144,10290) |
| Philippines | 599000 | 12.3 (8.4,16.7) | 30.0 (7.5,59.8) | 179905 (44705,358595) |
| Russian Federation | 73000 | 1.8 (1.1, 2.6) | 5.9 (1.0,18.9) | 4315 (727,13774) |
| Sierra Leone | 23000 | 10.3 (6.4, 14.9) | 26.4 (5.5,57.0) | 6082 (1263,13114) |
| South Africa | 360000 | 4.7 (3.2,6.4) | 14.1 (3.0,36.4) | 50730 (10704,130904) |
| Thailand | 105000 | 8.6 (5.6,12.3) | 23.1 (4.9,52.1) | 24239 (5088,54707) |
| URT | 137000 | 11.0 (7.5,15.1) | 27.7 (6.6,57.3) | 38004 (9088,78524) |
| Viet Nam | 170000 | 17.7 (13.6,22.1) | 38.2 (12.1,66.4) | 64914 (20942,112843) |
| Zambia | 59000 | 11.2 (7.0,16.0) | 28.1 (6.1, 58.9) | 16581 (3582,34724) |
| Zimbabwe | 29000 | 7.9 (5.0, 11.4) | 21.6 (4.3,50.3) | 6268(1256, 14583) |

\*Source for age standardised BMI<18.5 kg/m<sup>2</sup> in both sexes reported in Global Health Observatory
database on BMI (accessible at https://www.who.int/data/gho/data/themes/topics/topic-details/GHO/body-mass-index);
≠ Revised RR as per the latest evidence (supplementary material for the rationale)<sup>10</sup>
Abbreviations: CAR= Central African Republic; DPRK=Democratic People's Republic of Korea;
DRC=Democratic Republic of the Congo; PAF= Population Attributable Fraction; RR= Relative Risk;
PNG=Papua New Guinea; URT=United Republic of Tanzania

Tables 2 and 3 also show the impact of this corrected and revised estimate on the number of cases attributable to undernutrition in India, the country with the highest burden of TB as well as prevalence of undernutrition. According to current estimates a quarter of the cases in India may be related to undernutrition (0.67 millions). Using a correction of PAF based on prevalence of undernutrition, this figure rises to one-third of incident cases (0.9 million), while the estimated PAF using a revised RR is nearly half of incident cases (1.2 millions). Figure 3 describes the TB incidence in all 30 high burden countries, the current, corrected and the revised figures (in thousands).

**Figure 3:**
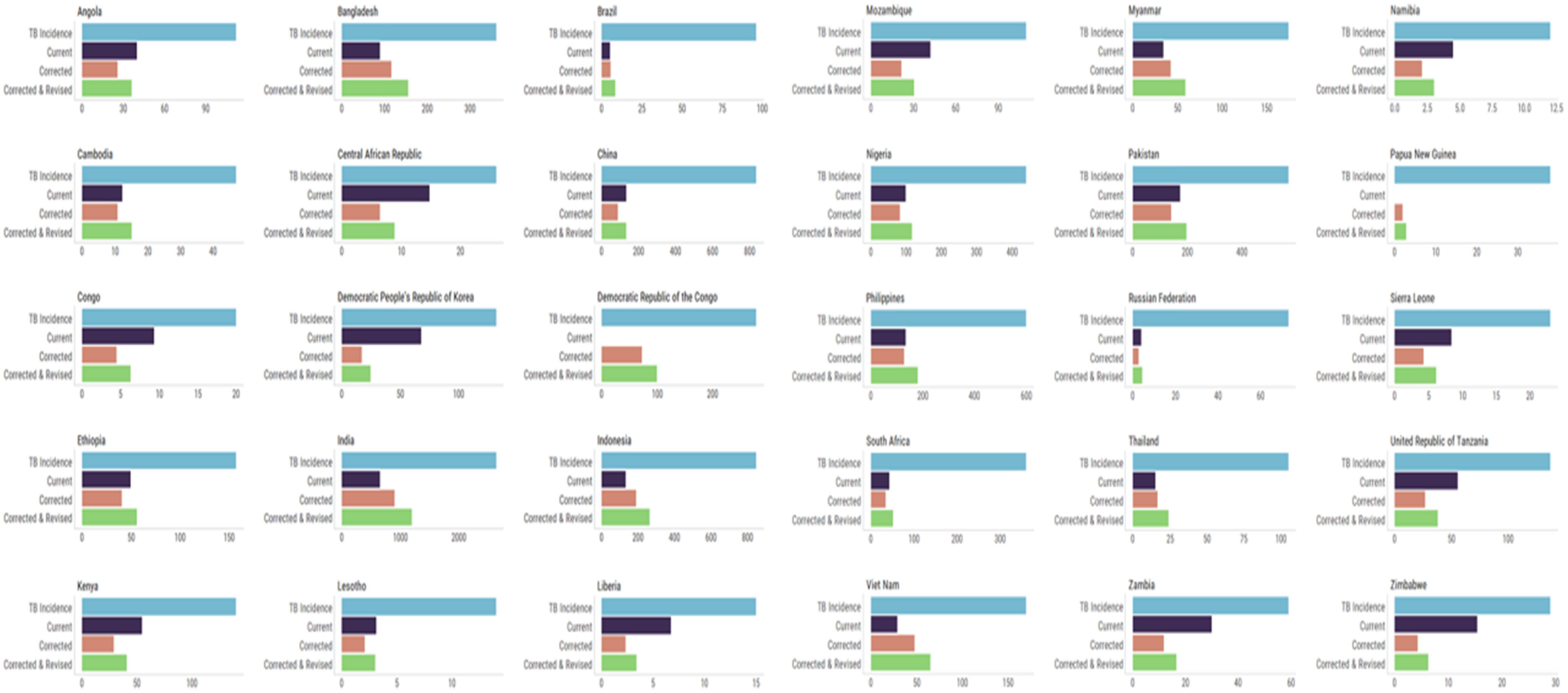
Number of cases of tuberculosis (in thousands) based on current, corrected and revised population attributable fraction (PAF) related to undernourishment and undernutrition in 30 high TB burden countries. **TB incidence**: Reported incidence in the Global TB Report 2020; **Current:** Number of cases of TB based on Prevalence of Undernourishment by Food and Agriculture Organization and relative risk (RR) of 3.2 as per the Global TB Report 2020; **Corrected:** Number of cases of TB based on prevalence of undernutrition (Global Health Observatory) and RR of 3.2; **Revised:** Number of cases of TB based on prevalence of undernutrition (Global Health Observatory figures) and revised RR of 4.49; In the absence of data on the Prevalence of Undernourishment, current PAFs for undernourishment were not estimated for the Demographic Republic of the Congo and Papua New Guinea.

## Discussion

We noted and addressed a methodological issue affecting the validity of the 2010 and 2020 estimates attributable to undernutrition and undernourishment, respectively. These estimates appear to have interpreted undernutrition and undernourishment as interchangeable terms. The PAF of undernutrition in the 2010 estimate thus used the prevalence of undernourishment (rather than prevalence of low body mass index or undernutrition). The recent estimate of PAF of undernourishment was estimated using the relative risk of undernutrition, since there is no relative risk for TB available for the exposure of undernourishment.

We present for the first time, methodologically corrected estimates of the PAF of TB related to undernutrition, using both the prevalence and relative risk of the same risk factor of undernutrition. We also reported the number of cases attributable to undernutrition in high TB burden countries. Overall, the median PAF of undernutrition was 19.7% compared to the median PAF of 28.3% of undernourishment. In 30 high TB burden countries, 24% or 2.07 million out of 8.62 million cases were attributable to undernutrition. In most Asian countries like India, PoU is less than prevalence of undernutrition and it underestimates the numbers who are underweight. On the other hand, the FAO estimates of undernourishment overestimate the proportion of people who are underweight in Africa. Overall undernutrition assumes an even greater imperative in the light of the COVID-19 pandemic, as food prices soar, as estimates suggest a doubling of the number of people experiencing acute hunger with serious implications for the TB epidemic [1,24].

After correction of the estimates, although the overall proportion of cases attributable to undernutrition increased only marginally, undernutrition emerged as the leading risk factor for TB in Asia. For example, the PAF based on PoU for India was 24.8% while that based on prevalence of undernutrition was 34%, a substantial difference for a country with the highest TB burden in the world. Using the correct estimates of prevalence of undernutrition did not alter the global burden of cases attributable to undernutrition significantly (1.96 million to 2.07 million). However it helped identify specific high TB burden countries like India and other Asian countries with low prevalence of HIV, where undernutrition is the major risk factor for TB. These are the countries where TB prevention efforts may benefit significantly from public health measures to reduce the levels of adult undernutrition. Shifting the population distribution of BMI in adults towards a desirable BMI level could have a marked impact on TB incidence, as indicated by a meta-analysis and modeling studies [7, 19].

On replacing the RR of 3.2 with 4.49 for the revised estimates, the PAF of undernutrition increased to a median PAF of 28%. As a result, the number of cases attributable to undernutrition increases to 2.84 million of the total 8.62 million (32.9%) incident TB cases. This suggests that one-third of TB cases in 30 high TB burden countries could be attributable to undernutrition, and therefore could be potentially eliminated if undernutrition was addressed. This represents an absolute increase of 0.88 million cases or a relative increase of nearly 45% of incident cases attributable to undernutrition, compared to current estimates.

### Strengths and limitations

Our study provides a valid estimate of the impact of undernutrition on the TB epidemic in 30 high TB burden countries. The use of the metric of undernutrition rather than undernourishment has implications for various National TB control programs. The prevalence of undernourishment determined by FAO does not allow disaggregated analysis to identify specific vulnerable populations in a country [25]. The periodic demographic and health surveys which generate prevalence of undernutrition in children and adults however can highlight especially vulnerable populations. Our higher revised estimate of RR for undernutrition is based on results from a cohort which had adjusted for the effects of confounders [10], and from a recent cohort from a high burden country with low HIV prevalence [20].

Our study has certain limitations. PAF is a static measure and some of the impact of addressing undernutrition is still not captured in these estimates [26]. In the case of transmissible diseases like TB, the prevention of progression to active disease by improved nutrition will also decrease the onward transmission of infection and prevent secondary cases. Measures like the transmission PAF may capture this impact [27]. We have not accounted for the interactions between undernutrition and other risk factors like HIV, diabetes, where a low BMI is also a risk factor for active TB [28], and the risk of TB is significantly higher for patients with diabetes who have a low BMI [29]. In fact comparatively the lowest risk was observed in obese patients with diabetes [29]. The association between undernutrition and incident TB in these estimates is an ecologic association and it cannot be confirmed whether the incident cases are actually occurring in those who have undernutrition. The estimates of low BMI in countries do not entirely capture the burden of undernutrition in the community. In India, for example, according to the National Family Health Survey-4, the prevalence of low BMI in adults is 23.6%, but a much higher proportion of children suffer from undernutrition; 38% of children under five had stunting (low height-for-age) and 36% were underweight (low weight-for-age) [30]. The wide confidence intervals around the RR attributable to undernutrition are due to paucity of cohort studies in populations with undernutrition. There is a need for larger cohort studies to estimate the RR of undernutrition which may be significantly higher than the 3.2 being currently used.

## Data Availability

All data produced in the present work are contained in the manuscript

## Conclusion

The estimation of PAF related to undernutrition rather than undernourishment is methodologically valid and operationally relevant and should be the basis of future such estimations. We report that with corrected and revised estimates of PAF of undernutrition, 2.07-2.84 million (24%-33%) of the 8.62 million TB cases in high burden countries may be attributable to undernutrition. The PAF of undernutrition at sub-national levels can provide actionable data for TB control efforts by identifying specific vulnerable populations with higher prevalence and PAFs of undernutrition. Undernutrition is the leading driver of TB globally and in the highest TB burden countries of Asia, and should be the focus of multi-sectoral efforts to reduce TB incidence in line with the goals of the END TB strategy.

## Contributors

AB, MB AnB and AK conceptualized the study, AB and AnB did the data curation, formal analysis and visualization. AB and MB wrote the original draft; all authors were involved in reviewing and editing the versions thereafter.

## Data sharing

All empirical data used in this study are made either publicly available, in the supplementary material or available from public domain.

## Acknowledgements

Authors acknowledge the data visualization and graphs done by Mr Aman Bhargava, a student of Bachelors in Design at Srishti Manipal Institute of Art, Design and Technology, Bangalore

